# Serum Troponin T levels as a therapy response marker in SOD1-mediated ALS

**DOI:** 10.1101/2025.02.13.25322198

**Authors:** Sarah Bernsen, Rachel Fabian, Yasemin Koc, Peggy Schumann, Peter Körtvélyessy, Sergio Castro-Gomez, Thomas Meyer, Patrick Weydt

**Affiliations:** Department of Neuromuscular Diseases, Center for Neurology, University Hospital Bonn, Bonn, 53127, Germany; Deutsches Zentrum für Neurodegenerative Erkrankungen, Research Site Bonn, Bonn, Germany; Department of Neurology, Center for ALS and Other Motor Neuron Disorders, Charité – Universitätsmedizin Berlin, Corporate Member of Freie Universität Berlin, Humboldt-Universität zu Berlin and Berlin Institute of Health, Berlin, Germany; Ambulanzpartner Soziotechnologie APST GmbH, Berlin, Germany; Deutsches Zentrum für Neurodegenerative Erkrankungen, Research Site Magdeburg, Magdeburg, Germany; Department of Parkinson, Sleep and Movement Disorders, Center for Neurology, University Hospital Bonn, Germany; Institute of Physiology II, University Hospital Bonn, Bonn, Germany

## Abstract

**Introduction:** Serum cardiac troponin T (cTnT) levels are elevated in the majority of amyotrophic lateral sclerosis (ALS) patients and increase over time. Neurofilament light chain (NfL) is an established therapy response biomarker in ALS as superoxide dismutase 1 (SOD1)-ALS patients treated with the antisense oligonucleotide tofersen show a decrease in NfL. In this study we assess the course of cTnT levels in SOD1-ALS at baseline and during tofersen treatment.

**Methods:** Serum cTnT was analyzed at baseline and during tofersen treatment in 23 SOD1-ALS patients at two specialized ALS centers in Germany and compared to a control cohort of 74 ALS patients without SOD1-mutations and not treated with tofersen.

**Results:** cTnT levels increased in the control ALS-cohort over time (p<0.0001) but not in the tofersen group (p=0.36). CK, and CK-MB levels did not show relevant changes over time. The median monthly increase of cTnT was 0.045 points (IQR 0.02-0.08) in the control ALS cohort and 0.01 points (IQR −0.01-0.03) in the tofersen group (p=0.0013). The fold change in cTnT levels of the tofersen-treated cohort (median 1.2; IQR 0.77-1.59) was significantly less and even showed a reduction in some patients compared to the control group (median 1.89; IQR 1.35-2.75) (p=0.0003).

**Discussion:** In this study, we find a response signal of cTnT to tofersen treatment, which supports the value of cTnT as an independent biomarker in ALS. These results contribute to the notion that cTnT may provide additional value as a progression and treatment response biomarker in ALS complementary to NfL and warrant further investigations.

**Graphical Abstract:** 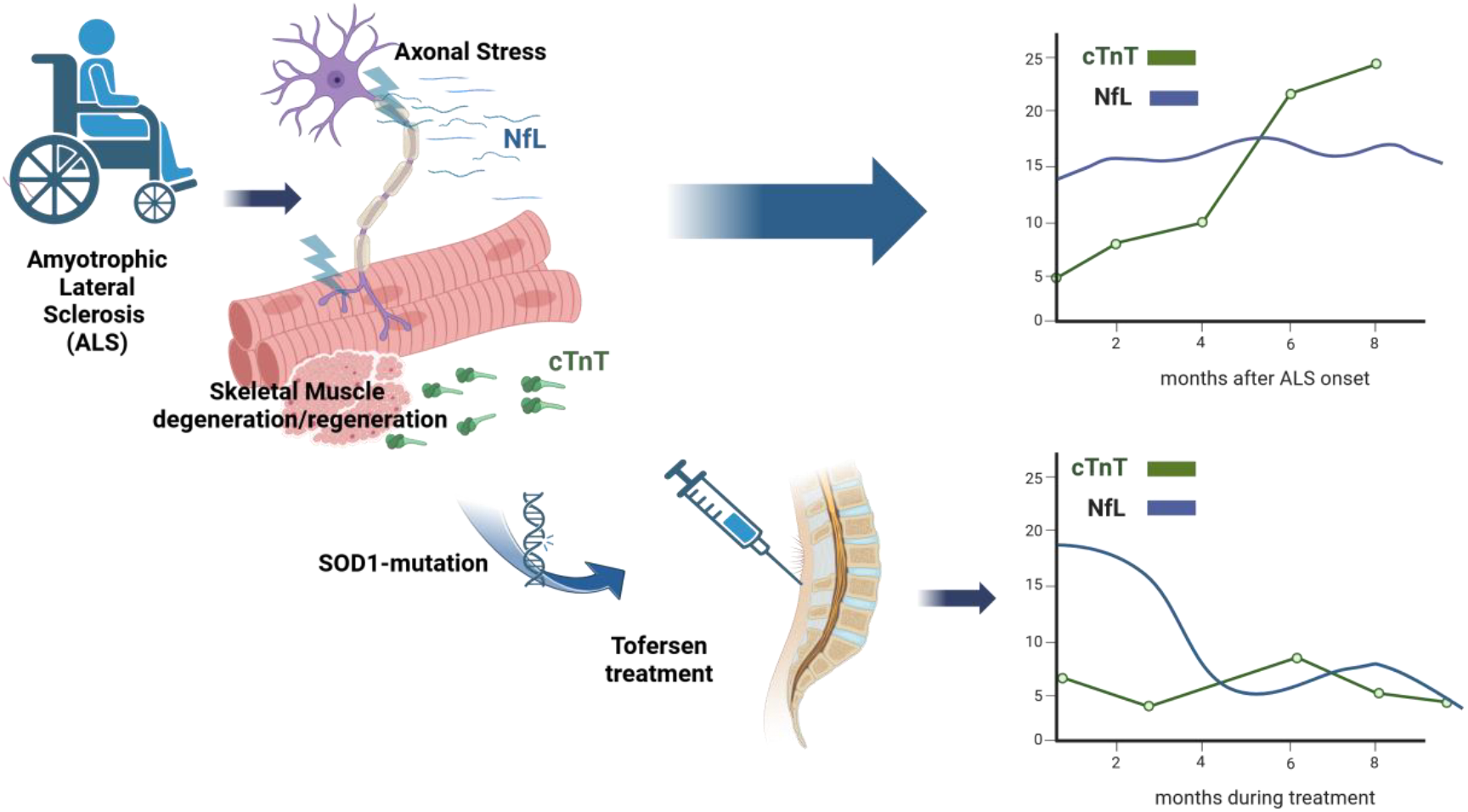

## Introduction

Amyotrophic lateral sclerosis (ALS) is a fatal neuromuscular disorder marked by degeneration of upper and lower motor neurons resulting in progressive muscle paralysis^1^. Death from respiratory failure occurs typically within three to five years from symptom onset. Recently the antisense oligonucleotide tofersen was approved as a disease-modifying drug for the small subset of ALS patients carrying a pathogenic superoxiddismutase1-(SOD1) gene mutation^2^. In these patients neurofilament light chain (NfL) levels in serum and cerebrospinal fluid (CSF), markers of neuroaxonal stress, are elevated and drop after treatment initiation^3^.

Serum cardiac troponin T (cTnT) levels are chronically elevated in nearly two thirds of the ALS patients^4^ and closely correlate with disease severity and progression^5–7^. Of note, cTnT levels are independent from NfL levels and their response to therapeutic interventions is unknown. Unlike other serum markers such as NfL, creatine kinase (CK) or CK-MB, cTnT levels increase over time^6,7^.

New disease modifying interventions offer the opportunity to investigate the potential of cTnT as a therapy response marker. Here, we analyzed the temporal dynamics of cTnT in ALS-SOD1-patients treated with tofersen.

## Methods

### Study Design

This study is a retrospective analysis using clinical routine data (Bonn) and data from the “APST-registry study”, and “NfL-ALS”-study in Berlin.

### Subject Samples

Tofersen is now standard of care for SOD1-ALS, so placebo-controlled trials for these patients are no longer ethically feasible. Thus, we analyzed demographic, clinical information, and serum samples of SOD1-ALS patients treated with tofersen for at least 6 treatment cycles at two German ALS clinics in Bonn and Berlin, and compared them to ALS patients without SOD1 mutations that were not treated with tofersen.

Data for the tofersen treatment group (N=23) were collected in the four-week treatment intervals between June 2022 and July 2024 in Berlin and Bonn. As the laboratory measurements and clinical data in Bonn of the treatment and control group were part of the routine clinical work-up and retrospectively analyzed, no formal consent was needed according to our institutional ethics review board (Ethics Board decision letter 324/20, Bonn).

At the ALS clinic Berlin data (demographics, ALSFRS-R, clinical characteristics, NfL, cTnT, CK, and CK-MB) were obtained via the APST registry and its “NfL-ALS” sub-study.

For the longitudinal data of the ALS patients without SOD1-mutation (and thus tofersen-naïve), we included 74 patients from the ALS clinic Bonn who were followed up after the first visit for at least 4 months and had ≥3 cTnT measurements available between January 2019 and August 2023.

### Laboratory Markers

#### Bonn

All measurements were performed at fully accredited commercial laboratories as described previously^6^; CK and CK-MB (University Hospital Bonn central laboratory), cardiac cTnT (Labor Volkmann, Karlsruhe), serum NfL (University Medical Center of Ulm, Germany).

#### Berlin

cTnT, CK, and CK-MB concentrations were measured at the Labor Berlin – Charité Vivantes GmbH. Serum NfL concentrations were analyzed at the ALS center in Berlin, described previously^3^.

### Variables

Clinical data included sex, BMI, age at symptom onset, age at treatment onset (SOD1 cohort) or age at first visit (untreated ALS cohort), and disease duration. Symptom onset was defined as the date (in month and year) of the onset of motor functional deficits, as captured by the ALS-Functional Rating Scale Revised (ALSFRS-R)^8^. Progression rate (PR) (points of ALSFRS-R lost per month) was obtained and further classified in patients with slower (<0.5 ALSFRS-R/month), intermediate (≥0.5 and ≤1.0 ALSFRS-R/month), and faster (>1.0 ALSFRS-R/month) progression^9^.

### Statistical Analysis

Statistical description and analyses were performed using GraphPad Prism 10 (GraphPad, San Diego, California, USA).

cTnT, NfL, CK and CK-MB presented a skewed distribution, so data are displayed as medians and nonparametric tests were used. Comparisons between groups were calculated with the Chi-Square-test for nominal variables and non-parametric the Mann–Whitney U test for non-normally distributed variables. To analyze changes over time between baseline and last visit the Wilcoxon matched pairs signed rank test was applied. We used Spearman’s rank correlation coefficients to measure the correlation between cTnT, CK, CK-MB and NfL. Because of missing values, cTnT, CK, and CK-MB samples were analyzed by a mixed-effects model instead of a repeated-measures analysis of variance. Data were log-transformed for the mixed effects-analysis to approximate normal distribution. Missing values were not replaced. *P* ≤ 0.05 was considered significant.

## Results

### Patient cohort and clinical characteristics

Clinical and demographic characteristics of the tofersen treated SOD1-ALS patients (N=23) and the control ALS group (N=74) are summarized in Table 1.

**Table 1:** Demographic and clinical characteristics of the two cohorts.

|  | ALS, tofersen treated | ALS, control | p-value for difference |
| --- | --- | --- | --- |
| Number of Patients | N=23 | N=74 |  |
| Female, n (%) | 11 (48%) | 25 (34%) | 0.2234 |
| Age at onset, median (IQR), years | 51.9 (38.7-60.6) | 61 (56.05-71.53), (74) | <0.0001 |
| Age at first visit, median (IQR), years | 58.8 (49.8-65) | 62.8 (58.18-73.2) | 0.0043 |
| BMI at first visit (kg/m<sup>2</sup>), median (IQR) | 24.49 (22.1-27.55) | 24.85 (22.4-27.75) | 0.7064 |
| Type of onset, n (%) | Bulbar: 1 (4%)<br>Spinal: 22 (96%) | Bulbar: 23 (31%)<br>Spinal: 51 (69%) | 0.0095 |
| Genetic mutation other than SOD1 | - | 14 (19%) |  |
| ALSFERS-R at first visit, median (IQR) | 34 (28.5-41.25) | 36 (32-41) | 0.2841 |
| ALSFERS-R at last visit, median (IQR) | 34 (24-42) | 24 (18-29) | 0.0013 |
| Tofersen treatments, median, IQR | 16 (7-20) | - | - |
| Months of follow-up, median (IQR) | 14 (6-22) | 12 (9-19,25) | 0.6808 |
| Onset to first visit, months, median (IQR) | 43 (22-80) | 14 (7.8-124) | <0.0001 |
| PR (onset to first visit) (median, IQR) | 0.31 (0.15-0.58) | 0.64 (0.33-1.1) | 0.0014 |
| PR (onset to last visit) (median, IQR) | 0.21 (0.11-0.49) | 0.78 (0.55-1.36) | <0.0001 |
| **PR Classification** | **N=22** | **N=72** |  |
| Slow (<0.5), n (%) | 16 (73%) | 24 (33%) | 0.0047 |
| Intermediate (≥0.5 and ≤1.0), n (%) | 3 (14%) | 23 (32%) |  |
| Fast (>1.0), n (%) | 3 (14%) | 25 (34%) |  |

ALS=Amyotrophic Lateral Sclerosis; ALSFRS-R=Amyotrophic Lateral Sclerosis Functional Rating Scale Revised; BMI= Body Mass Index; IQR= Interquartile Range; N/n= Number; PR=Progression Rate; SOD1=Superoxiddismutase1

### Serum biomarkers

At first sampling, the tofersen-treated cohort presented with higher CK-MB levels (p=0.0013) and lower NfL-levels (p<0.0001) in comparison to the non-SOD1 ALS patients. All biomarker results are described in Table 2.

**Table 2:** Serum biomarker characteristics of the two cohorts.

|  | ALS, tofersen treated | ALS, control | p-value for difference |
| --- | --- | --- | --- |
| First cTnT in serum (ng/l)<br>(median, IQR) | 20.8 (11-42) | 20.7 (11.58-32.75) | 0.7956 |
| Last cTnT in serum (ng/l)<br>(median, IQR) | 19 (12-57) | 35.1 (19.65-64.2) | 0.128 |
| First cTnT in Serum<br>> cut-off * | 15 (65%) | 54 (73%) | 0.4734 |
| First NfL in serum (ng/l)<br>(median, IQR) | 57.8 (21.47-78) | 113.5 (91.25-171) | <0.0001 |
| Last NfL in serum (ng/l)<br>(median, IQR) | 22.7 (9.6-54.6) | 135 (70-174) | <0.0001 |
| First CK in serum (U/l)<br>(median, IQR) | 208 (119.5-443.5) | 200 (116.5-347) | 0.5388 |
| Last CK in serum (U/l)<br>(median, IQR) | 190 (117-375) | 122 (77.5-239.5) | 0.0655 |
| First CK-MB in serum (U/l)<br>(median, IQR) | 14.9 (8.1-20.4) | 7.5 (3.8-12.6) | 0.0013 |
| Last CK-MB in serum (U/l)<br>(median, IQR) | 12.9 (6.2-17.1) | 6.3 (4.1-12.5) | 0.008 |
| **<u>Slow progressors</u> \*\*** |  |  |  |
| First cTnT in serum (ng/l)<br>(median, IQR) | 17 (11.5-38.5) | 25.7 (10.3-51.8) | 0.6162 |
| Last cTnT in serum (ng/l)<br>(median, IQR) | 18 (10.5-53.6) | 56.9 (14.7-75) | 0.0695 |
| First NfL in serum (ng/l)<br>(median, IQR) | 50.55 (24.09-70.9) | 103.5 (59-141.3) | 0.064 |
| Last NfL in serum (ng/l)<br>(median, IQR) | 22.4 (11.2-39) | 77 (40-169.5) | 0.0006 |

CK= Creatine Kinase, cTnT=cardiac Troponin T, IQR=interquartile range, NfL=Neurofilament light chain, PR=Progression Rate
\*cut off >14 ng/l,
\*\* PR<0.5 ALSFRS-R/month

As expected, cTnT levels in the control-cohort increased over time (p<0.0001). In the tofersen-treated patients, in contrast, the cTnT levels remained unchanged (p=0.36). CK in the tofersen cohort, and CK-MB levels in both groups (first vs last sampling) did not show a significant change. Interestingly, CK levels showed a significant decrease over time only in the control group (p=0.008) (Fig. 1A and 1B).

**Figure 1.**
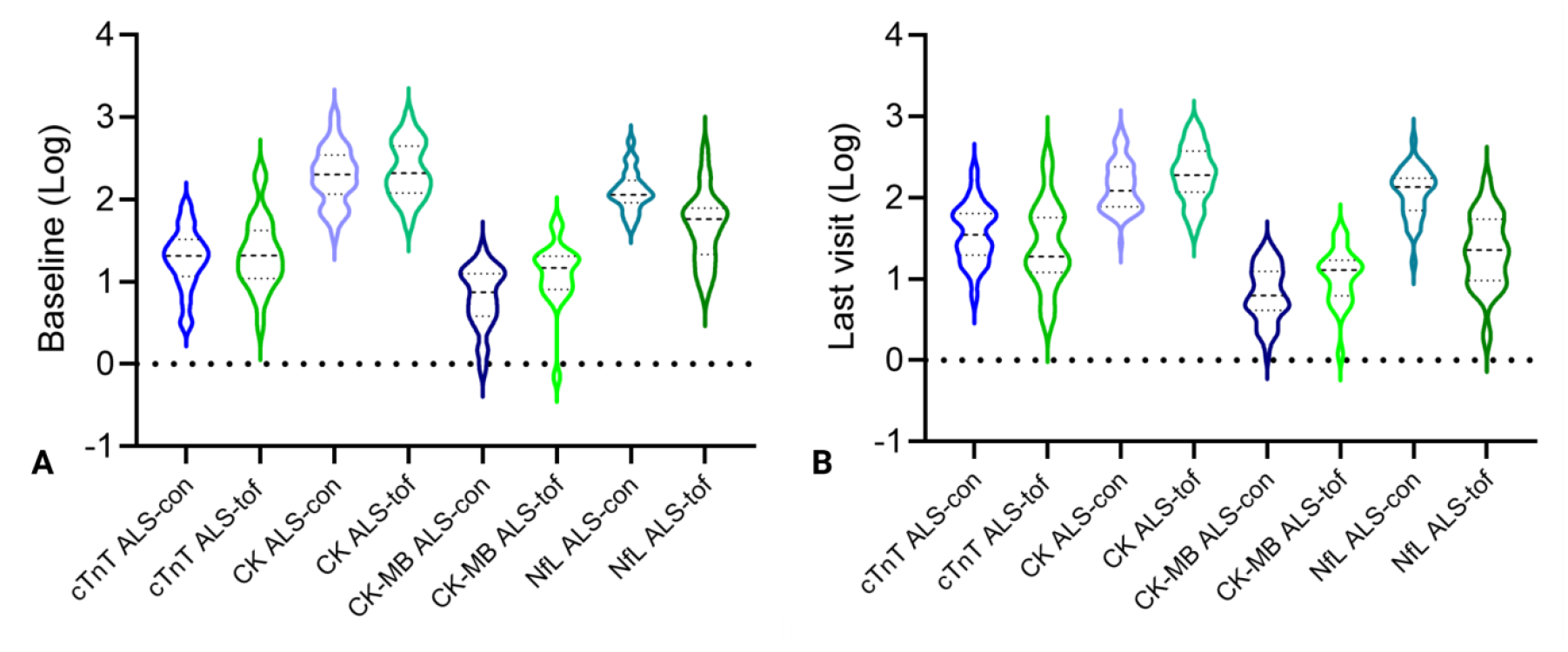
**A** Baseline levels (log) of cTnT, CK, CK-MB and NfL of SOD1-ALS patients treated with tofersen (ALS-tof) and control ALS-patients (ALS-con) without tofersen treatment. Violin plot represents the actual distribution median (dashed line) and quartiles (dotted line). **B** cTnT, CK, CK-MB and NfL values (log) at last visit of SOD1-ALS patients treated with tofersen (ALS-tof) and control ALS-patients (ALS-con) without tofersen treatment. Violin plot represents the actual distribution median (dashed line) and quartiles (dotted line).

The compound monthly growth rate (CMGR) for cTnT was 0.045 points (IQR 0.02-0.08) in the ALS control cohort and 0.01 points (IQR −0.01-0.03) in the tofersen-patients (p=0.0013). In the treatment group cTnT correlated moderately with CK-MB, (p=0.047, r=0.45), but not with NfL and CK. In the ALS patients without tofersen treatment cTnT correlated strongly with CK-MB (p=0.0001, r=0.64) and CK (p=0.0001; r=0.49), but not with NfL levels.

Moreover, we measured cTnT repeatedly over time during 19 months. A mixed-effects model for repeated measures showed a significant interaction of time and treatment between both groups for cTnT (p=0.0013) (Fig. 1C), but not for CK (p=0.5734) or CK-MB (p=0.4344) levels (Fig.2A).

The fold-change in cTnT levels of the tofersen-cohort (median 1.2; IQR 0.77-1.59) was significantly less and even showed a reduction in some patients compared to the control ALS-patients (median 1.89; IQR 1.35-2.75) (p=0.0003). This difference remained consistently statistically significant if stratified for the slow PR subgroup (p=0.009). There was no difference in fold change of CK and CK-MB (data not shown). The fold change in NfL levels decreased in the SOD1-patients as expected (median 0.56; IQR 0.32-0.85) compared to the control group (median 1.01; IQR 0.75-1.25) (p=0.0005) (Fig. 2B).

**Fig 2.**
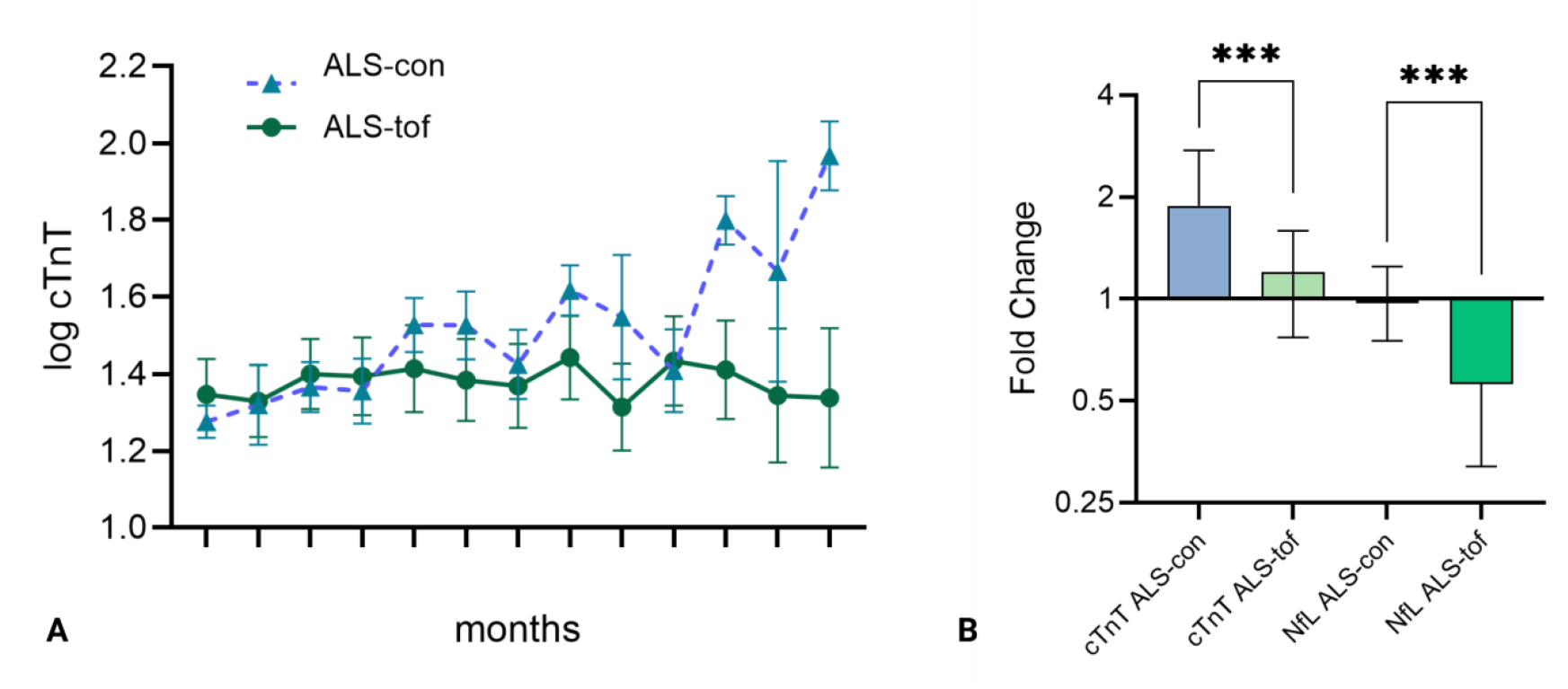
**A** cTnT changes over time (mean, SEM) of SOD1-ALS patients treated with tofersen (ALS-tof) and control ALS-patients (ALS-con) without tofersen treatment prior to first tofersen administration (ALS-tof) or first visit (ALS-con) up to 19 months compared with a mixed linear effects model for repeated measures (p=0,0013). **B** Fold Change of cTnT and NfL from SOD1-patients treated with tofersen (ALS-tof) compared with control ALS patients (ALS-con) without tofersen treatment (cTnT*** p=0,0003, NfL*** p=0,0005). Whiskers show IQR.

We then normalized the data by calculating the change of biomarker levels between the longest available interval as Δ divided by the elapsed time (days). SOD1- and control ALS-patients were significantly different in regard to Δ cTnT (p=0.0085), and Δ CK-MB (p=0.0085) but not Δ CK (p=0.82) (Fig 3A and 3B). Even stratified for the subgroup of the slow progressors the effect remained statistically significant for Δ cTnT (p=0.003).

**Fig 3.**
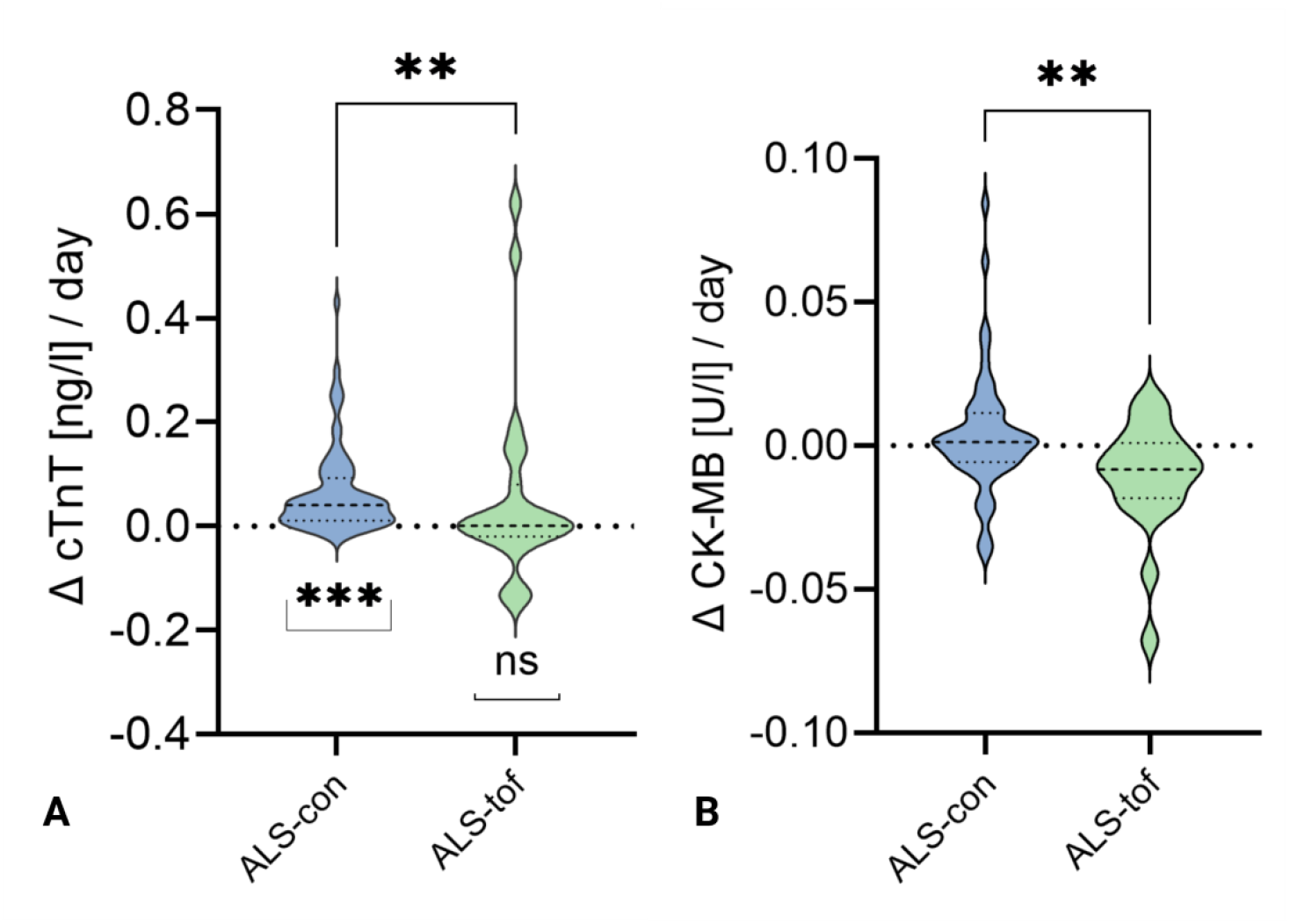
**A** Change in time (*Δ* cTnT/days) of cTnT levels of SOD1-patients with tofersen treatment (ALS-tof) and control ALS patients (ALS-con) without tofersen treatment between baseline and last visit. The two groups are compared with Mann–Whitney test (upper asterisks; p**=0,0085), the level of significance below each group is from Wilcoxon Signed Rank Test on *Δ* cTnT concentrations compared to the theoretical median of no change (p***<0,0001). Violin plot represents the actual distribution median (dashed line) and quartiles (dotted line). **B** Change in time (*Δ* CK-MB/days) of CK-MB levels of SOD1-patients with tofersen treatment (ALS-tof) and control ALS patients (ALS-con) without tofersen treatment between baseline and last visit. The two groups are compared with Mann–Whitney test (upper asterisks; p**=0,0085). Violin plot represents the actual distribution median (dashed line) and quartiles (dotted line).

## Discussion

In this study, we describe a response signal of serum cTnT levels to tofersen treatment in ALS patients. While there is a continuous increase of cTnT in ALS patients without tofersen treatment over time, the cTnT levels stabilize in tofersen treated ALS patients. cTnT shows a response to tofersen treatment similar to the previously observed effect of NfL in serum and CSF, thus suggesting it may have use as a complementary biomarker for monitoring therapeutic effect in ALS patients.

Serum NfL levels were lower in the tofersen-treated group at first sampling reflecting the overall lower PR in this group. This corresponds with the higher CK-MB baseline levels in this group, as those coincide with slower disease progression^10^.

In line with previous studies, CK and CK-MB showed no consistent changes over time^10^. The declining of CK levels over time in both ALS cohorts may reflect progressive loss of muscle mass. CK-MB, showing a closer correlation with cTnT than CK in both cohorts, only had a significant decrease in the tofersen-group when normalized for the elapsed time, suggesting a potential value as a marker of skeletal muscle involvement that has to be explored in the future^10,11^.

The results of the present study, highlight the potential of cTnT and CK-MB^12^ as markers of peripheral involvement in ALS. Interestingly, these peripheral, presumably muscle biomarkers show a dynamic response to a centrally acting therapy such as tofersen. The broad availability, dynamics, and temporal pattern of cTnT makes it unique and distinct from NfL and CK-MB, and suggests an immediate potential application in evaluating therapeutic effects.

Some of the limitations of our study are the small treatment group and the predominance of SOD1-ALS patients with slow progression rates. We have attempted to address this by stratification for the subgroup of slow progressors. A comparison with untreated SOD1-patients is not feasible, as all SOD1-patients in our clinics are on treatment with tofersen.

Our findings support the value of cTnT as an independent biomarker in ALS delivering information on therapy response and disease activity that has to be further investigated in future studies.

## Data Availability

All data produced in the present study are available upon reasonable request to the author

